# COVID-19 Rapid Diagnostic test results and their associations with certain factors among the residents of Balochistan

**DOI:** 10.1101/2020.12.07.20245076

**Authors:** Ehsan Larik, Muhammad Arif, Abid Saeed, Mirza Amir Baig, Zakir Hussain, Ambreen Chaudhary, Zeeshan Iqbal Baig, Zubair Bugti, Jan Inayat, Khair Muhammad, Muhammad Abdullah, Zubair Ahmed, Qurat-ul-ain, Aftab Kakar, Nasir Sheik

## Abstract

**Background:** This paper analyses any possible association of various factors like gender, last COVID-19 PCR test results, BCG Vaccination, Seasonal Flu vaccination, Occupation and confirmed case contact history with COVID-19 RDT results of the participants. COVID-19 will soon become endemic in Pakistan, the government should adopt COVID-19 RDT kits for trace, test and quarantine activities.

**Methodology:** Considering the availability of COVI-19 rapid diagnostic kits, **596** individuals all previously COVID-19 PCR tested were made part of this ***<u>cross sectional</u>*** study. Simple random sampling was used for the selection of study participants. The whole study was conducted during September and October 2020.

**Results:** The major findings of this study is clearly showing that the Positive Likely hood ratio of the COVID-19 RDT Kits (LR+) is well above 1; similarly the Negative Likely hood ratio is approaching 0.On the other hand the Sensitivity and Specificity 80% and 74% respectively. Similarly study found statistically significant association was between RDT out comes and Last PCR Test status, Occupation and Contact with COVID-19 positive individuals. While other variables like Gender, BCG Vaccination and history of seasonal flu vaccinations were found to have no significant associations with COVID-19 RDT Kit out comes.

**Conclusion:** Being the first study of its kind in Pakistan the major findings of this study are almost in line with the set hypothesis and objectives of this study and based on study findings it will be of high value to use COVID-19 RDT kits during mass screening especially during Test, Trace and Quarantine activities.

## Back Ground

The COVID-19 infections are characterized by highly nonspecific manifestations including respiratory symptoms, fever, cough, difficulty in breathing. These symptoms are also seen as clinical presentations of other virus-related diseases including influenza [1,2]. This poses a challenge in identifying those patients with COVID-19 from individuals with other respiratory diseases. Therefore, there is a need for a diagnostic test that is rapid, accurate and cost efficient that may be used at point-of-care to screen and confirm suspected cases. Early case detection has been proven to have a dramatic effect in controlling infectious disease outbreaks [3]. The current, WHO-recommended gold standard for the diagnosis of COVID-19 is the qualitative detection of SARS-CoV-2 virus nucleic acid via reverse transcription polymerase chain reaction (RT-PCR). RT-PCR is reported to have a sensitivity of 95% and a specificity of 100%; for every 100 COVID-19 positive patients, RT-PCR would have a falsely negative result in 5 patients [4]. The test, however, has limitations such as long turnaround times and complicated logistical operations that makes it infeasible as a rapid and simple field test option to screen and diagnose patients. Another proposed rapid, simple, and highly sensitive way to diagnose COVID-19 is through the qualitative detection of antibodies that are specific to SARS-CoV-2 instead of the direct detection and measurement of viral load through RT-PCR. Several studies have investigated the use of antibodies in the diagnosis of COVID-19 using ELISA [5, 6, 7] and lateral flow rapid test kits [8,9]. These studies show different sensitivity and specificity results, and different recommended timing of testing. Disparities between ELISA and lateral flow rapid test results may be due to the longer incubation time for ELISA compared to swift resolution for lateral flow tests, the slow kinetic dissociation rate of ELISA compared to a faster kinetic association rate in lateral flow tests, and the ELISA “capture” antibody and “detector” antibody designations may be reversed in lateral flow tests [10]. Thus, results from studies using ELISA and lateral flow tests should be analyzed separately.

A rapid point-of-care lateral flow immunoassay test product was developed intended for qualitative detection of IgM/IgG in human blood within 15 minutes. It has been designed to be a complementary aid in the diagnosis of patients suspected to have the COVID-19 infection. Limitations of the test include the following: 1) it does not directly confirm virus presence, instead, it provides serological evidence of recent infection, 2) it is not known if the test will cross-react with antibodies to other coronaviruses and flu viruses [8]. Further, studies in patients with Severe Acute Respiratory Syndrome (SARS), a disease also caused by a coronavirus, show that IgM is detectable as early as 3 to 6 days from symptom onset, while IgG is detectable after 8 days, with peak titers at 15 to 20 days [9,11]. This potentially limits the clinical and public health utility of antibody tests for the early diagnosis of coronavirus infections. Clinical trials that investigate the accuracy and safety of IgM/IgG rapid test kits for diagnosis of COVID-19 patients are still limited.

### Literature Review

This rapid review summarizes the available evidence on the accuracy and safety of lateral flow immunoassay (LFIA) IgM/IgG rapid test kits in diagnosing patients with COVID-19. Evidence on the accuracy and safety of the enzyme-linked immunosorbent assay (ELISA) test for COVID-19 are summarized in a separate rapid review.

Population: Symptomatic and asymptomatic COVID-19 patients and suspected COVID19 patients of any age, with any comorbidities, any severity

❖ Intervention: Antibody/Antigen test, IgM/IgG rapid test kit
❖ Comparator: Reverse Transcriptase Polymerase Chain Reaction (RT-PCR)
❖ Outcomes: Sensitivity, Specificity, Time to detection of antibodies
❖ Methods: randomized controlled trials (RCTs), non-randomized studies, cohort studies, case-control studies, cross-sectional studies.

After comprehensive search and appraisal, two (2) completed studies (Appendix 1) and one (1) ongoing trial (Appendix 2) on the accuracy of IgM/IgG antibody test kits for diagnosing COVID-19 were identified.Li et al., examined 525 blood samples of clinically positive (including PCR test) (n = 397) and clinically negative(n=128)patients to determine these sensitivity and specificity of the IgM /IgG rapid test kit [8].On the other hand,Yingetal.,investigated179 patients who were PCR positive (n=90) And PCR negative (n=89) comparing the over-all sensitivity and specificity of the antibody test kit and when done between day 0-7,day8-15, or day16 and beyond [9].In both studies, apositive test result was defined as detection of either or both IgM and IgG antibodies to SARS-CoV-2. A negative test result was defined as non-detection of any or both IgM and IgG antibodies to SARS-CoV-2 [8,9].The ongoing trial (NCT04316728) is designed to evaluate the clinical performance of IgM/IgG antibody test kits in the early diagnosis of COVID-19inhighriskpopulations.The study will serially test uninfected health-care workers and individuals with chronic conditions (n=200)and is expected to complete data collection by September2020.

### Critical Appraisal

Two studies provided direct evidence on the accuracy of lateral flow immunoassay (LFIA)IgM/IgG rapid test kits in diagnosing COVID-19 compared with PCR and clinical picture as the reference standard. Both studies did not adequately describe the methods used to validate the accuracy of the test kits. As such, it is difficult to ascertain whether or not safeguards to ensure a good estimate of the test kit’s diagnostic accuracy were in place; the absence of these safeguards would tend to result in an overestimate of the test kit’s diagnostic accuracy. As such, the results of these two studies need to be interpreted with caution.

### Accuracy Outcomes

The overall accuracy of the rapid test from the two identified studies are summarized below.

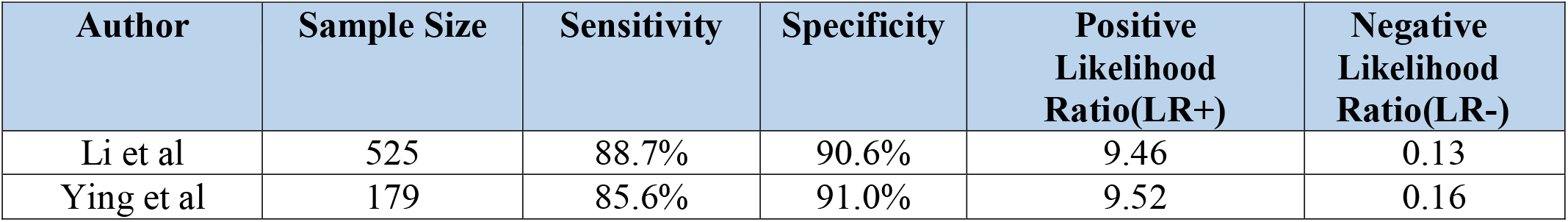

In majority of cases, Li et al was not able to determine the number of days from symptom onset to the time the blood sample for the rapid test was collected. However, in a subset of patients from one institution (n=58), the blood samples were collected at day 8 to 33 after symptom onset [8].

Ying et al reported the time from onset of illness to blood sample collection in 115 patients. The accuracy of the rapid test kit, stratified according to number of days of onset, is summarized below. The sensitivity (18.8%) of the rapid test was extremely low among those who had their blood samples collected within the first week of symptom onset [9].

Accuracy of IgM/IgG rapid test kits, stratified according to the number of days after onset of symptoms, Ying et al [9]

| Day test done | Sample Size | Sensitivity | Specificity | Positive Likelihood Ratio (LR+) | Negative Likelihood Ratio (LR-) |
| --- | --- | --- | --- | --- | --- |
| 0 to 7 days | 25 | 18.8% | 77.8% | 0.84 | 1.05 |
| 8 to 15 days | 8 | 100.0% | 50.0% | 2.0 | 0.17* |
| 16 days | 82 | 100.0% | 64.3% | 2.8 | 0.01* |

### Safety Outcomes

No adverse events were reported among the studies reviewed.

### Problem Statement

COVID 19 is a new disease not much is known about it and it would soon become endemic in Pakistan. Till 30^th^ August 2020, 1215 confirmed cases have been reported and local transmission is on its peak with 7 infected districts in Balochistan. The current cases of COVID-19 are iceberg of diseases, there may be many more cases in community.

### Rationale

The present standard for diagnosis of COVID-19 is through qualitative detection of COVID19 virus nucleic acid via reverse transcription polymerase chain reaction (RT-PCR). Due to long turnaround times and complicated logistical operations, a rapid and simple field test alternative is needed to diagnose and screen patients. There is delayed in results due to limited RT PCR Laboratory in Balochistan Province and its limited capacity to carry out daily PCR test. Even it was observed that results are declared after death of Positive case. As compared to RT-PCR, RDT is simple, cost effective and timely test to be conducted in field. As, Novel nature of disease, antibodies studies are conducted on limited level throughout Pakistan. Study is justifiable because so far no published study has ascertained the association of various known factors with immunity status using COVID-19 RDT results among suspected, confirmed and exposed individuals so that in the long run Possible COVID-19 exposure and encounter event could be made from individual’s history while performing Trace, test and Quarantine (TTQ) activities in the field till the development of a vaccine.

### Research Questions

**Q:** Is there any association between COVID-19 RDT out-comes and certain variable of interest like Gender, Last COVID-19 PCR results, BCG Vaccination, Seasonal Flu vaccination, Occupation and Close contact or living with confirmed case of COVID-19 in last 15-45 days ?

**Q:** Is COVID-19 RDT Kits suitable for diagnostic Purposes?

### Objectives

1. To ascertained the association of various known factors with immunity developed against COVID-19 using serological COVID-19 RDT for suspected, confirmed and exposed individuals.
2. To determine the Positive and Negative Likely hood Ratios for the COVID-19 RDT kits.
3. To ascertain clinical history based positive COVID-19 cases in the field while performing Test, Trace and Quarantine (TTQ) activities in the field till the development of a vaccine.

### Hypothesis

**H1**_**0**_: COVID-19 Serological Rapid Diagnostic test outcome has no association with Gender, Last COVID-19 PCR report, BCG vaccination, Seasonal flue vaccination, Occupation and Close contact or living history with confirmed case of COVID-19 in last 15-45 days.

**H1**_**a**_: COVID-19 Serological Rapid Diagnostic test outcome has association with Gender, Last COVID-19 PCR report, BCG vaccination, Seasonal flue vaccination, Occupation and Close contact or living history with confirmed case of COVID-19 in last 15-45 days.

**H2**_**0**_: There exists no association between COVID-19 RDT Kits out comes and COVID-19 Diagnosis.

**H2**_**a:**_ There exists an association between COVID-19 RDT Kits out comes and COVID-19 Diagnosis.

### Research Methodology

A ***Cross-sectional*** study was performed, considering the availability of COVID-19 Rapid Diagnostic test kits a total of **<u>596</u>** individuals were included in this study. All of these participants were selected by Simple random sampling after acquiring the “Complete name list” of all the individuals whom had done their PCR lab test in Fatimah Jinnah Chest Hospital Quetta. The study setting was District Quetta Balochistan,the whole population of Balochistan was considered as sample population. ***Every individual of any age and gender belonging to any district of Balochistan who had done his or her COVID-19 PCR test at PCR Lab Fatimah Jinnah Chest hospital Quetta from 1***^***st***^ ***January 2020 till 30***^***th***^ ***August 2020 were included in the study regardless of their PCR test reports***. Prior to data collection Ethical review board at Director General Health Services Balochistan was approached for the approval. Informed consents were acquired from each participant after briefing them. Data collection was done through interviews using structured questioner. Later on at the end of each interview a single fresh blood drop was acquired from every individual for COVID-19 Rapid Diagnostic test. The whole study was conducted during September and October 2020.

Descriptive statistics was used for age, sex, occupation, educations status, & Blood groups of the study participants while inferential statistics like Chi-square was used to assess various associations. SPSS Software −19 versions was used to analyze the data. Similarly Positive and Negative Likely hood ratios calculated using 2×2 table.

## Results

### Descriptive statistics

Out of 596 individuals 76.8% (n= 458) were males while 23.2% (n=138) were female participants as shown below:

**Fig-1:**
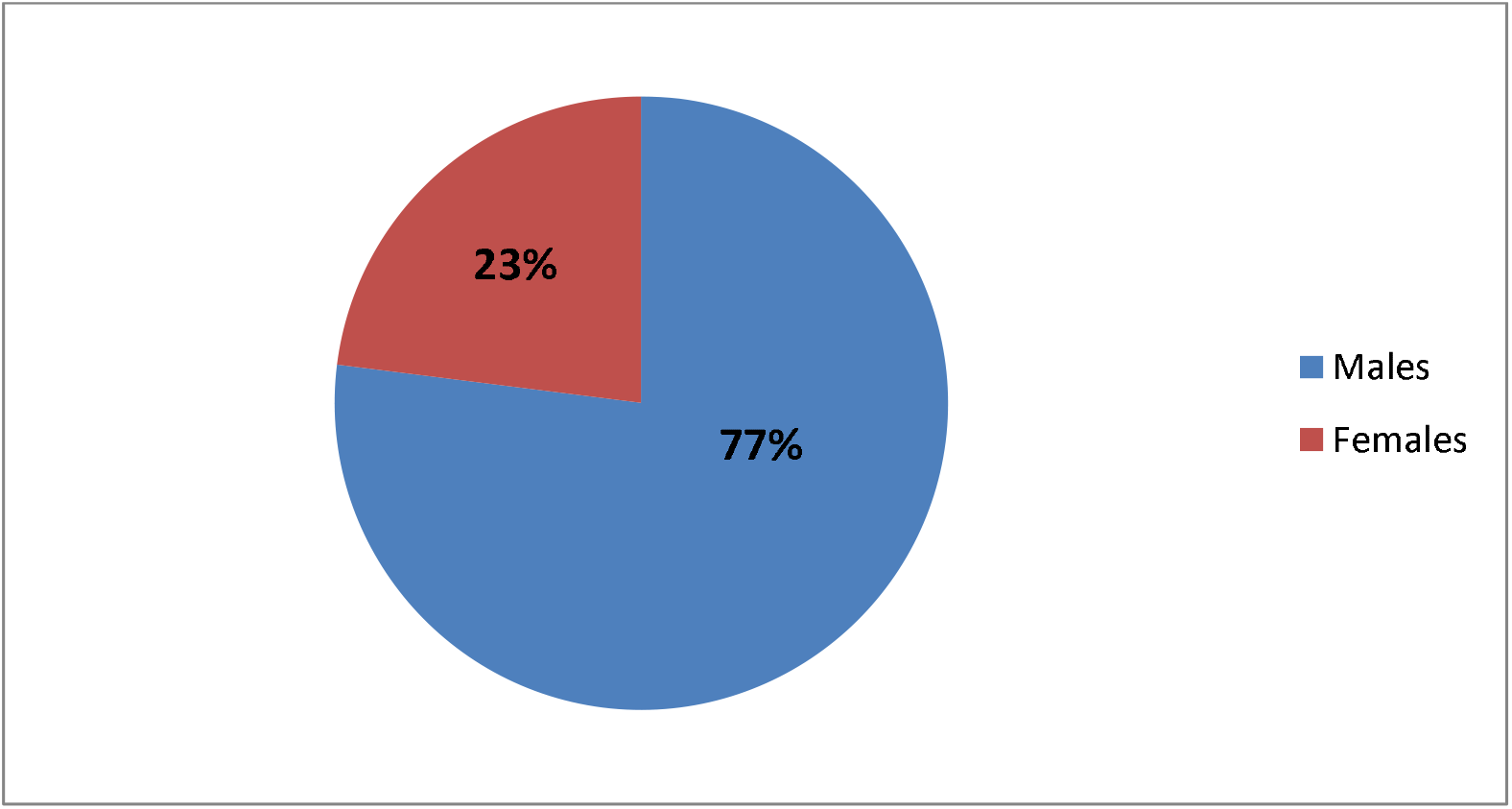
Gender wise Distribution of the Study Participants.

### Age Distribution

Most of the 596 participants in this study were young in their early thirties and forties. The age range of the study Participants ranged from 3 to 80 Years.

**Fig-2:**
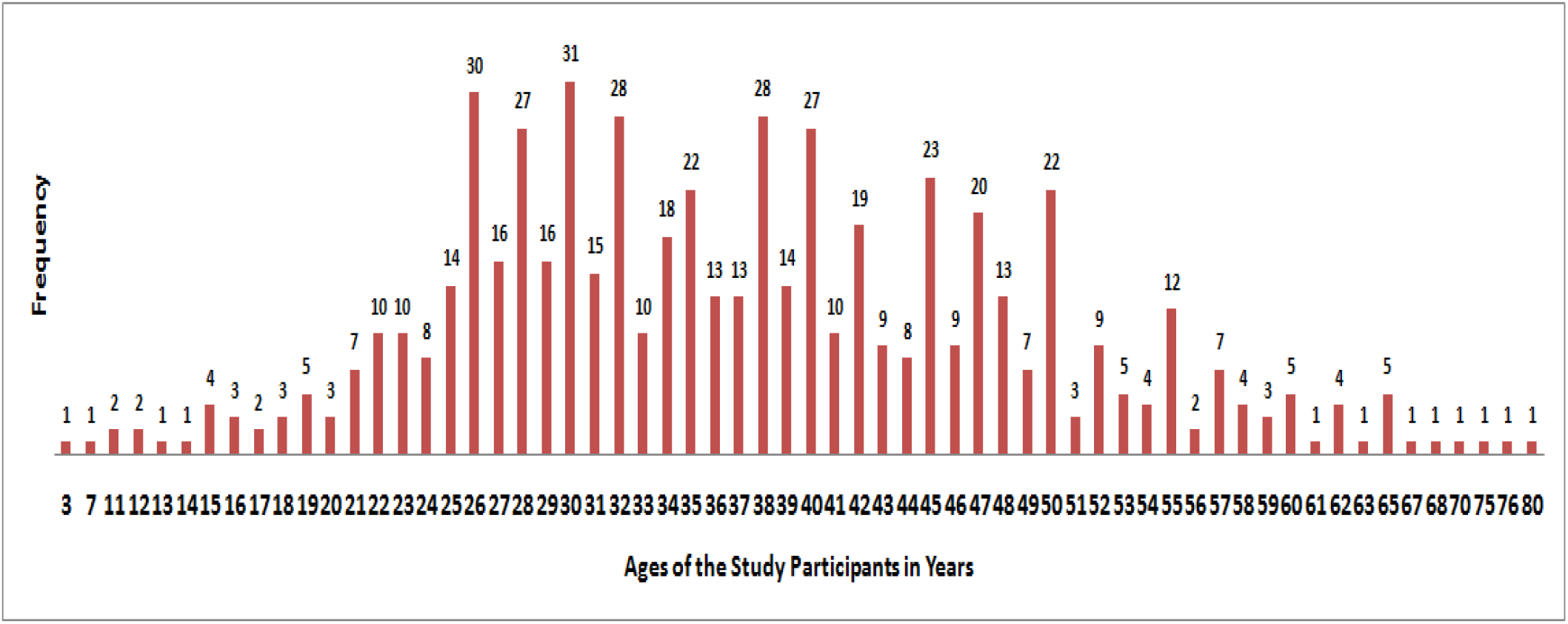
Age Distribution of the Study Participants.

### Education Level Of the Participants

Most of the study participants were educated,01% (n=06) were un educated,05% (n=30) had done primary education, 06% (n=38) had done Metric, 02% (n=13) of the study participants had done intermediate while 05% (n=28) had done graduation, 02% (n=09) had done Masters while non of the participants had Phd qualification :

**Fig-3:**
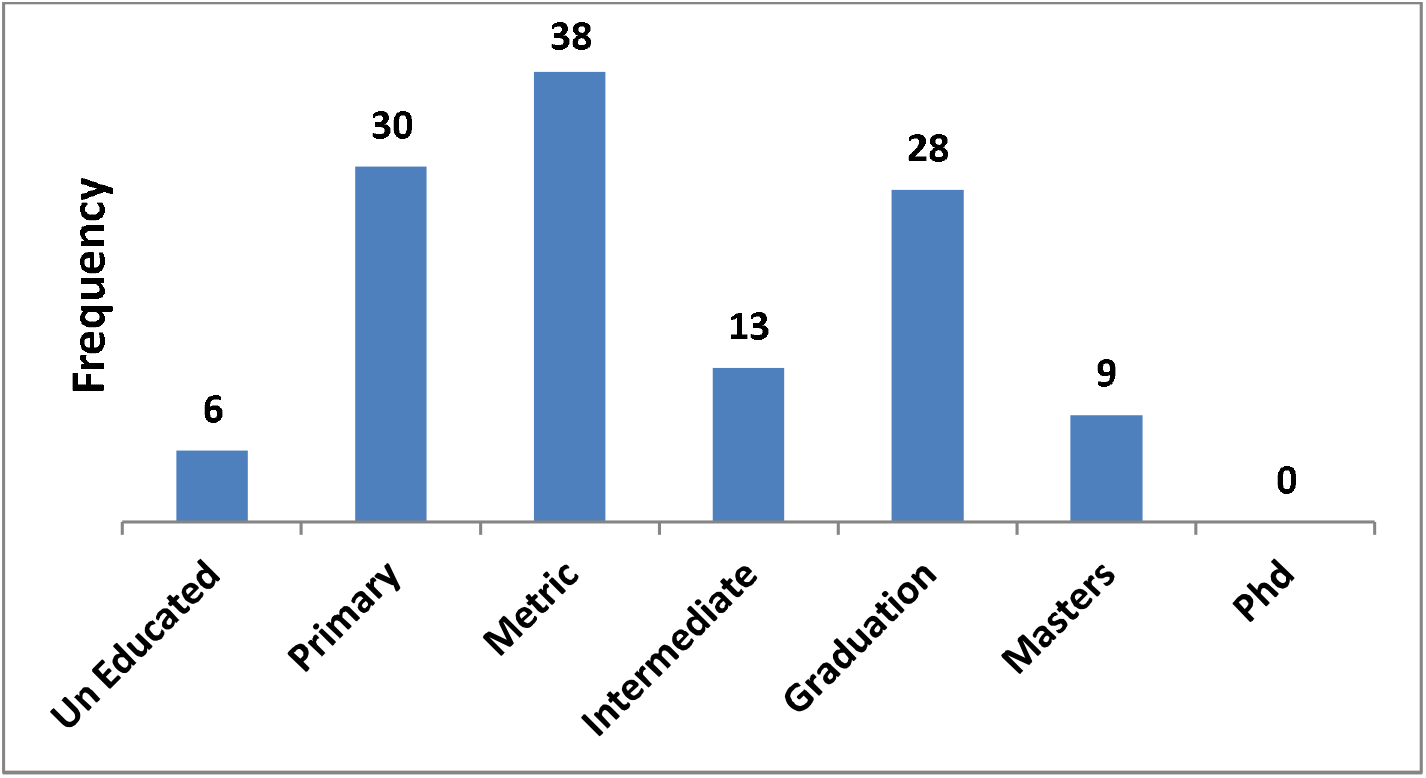
Education Level of the study Participants:

### Blood Groups of the Participants

Out of 596 Participants 13.6% (n= 81) had A+ Blood group, 1 % (n= 06) had A-Blood group, 13.8% (n=82) had B+ Blood group, 0.8 % (n=5) had B-Blood group, 3.7% (n=22) had AB+ blood group, while 0.2% (1) had AB-blood group similarly 14.8% (n=88) had O+ Blood group, 1.8% (n=11) had O-blood group, 50.3% (n=300) participants did not know about their blood groups when asked about.

**Fig-4:**
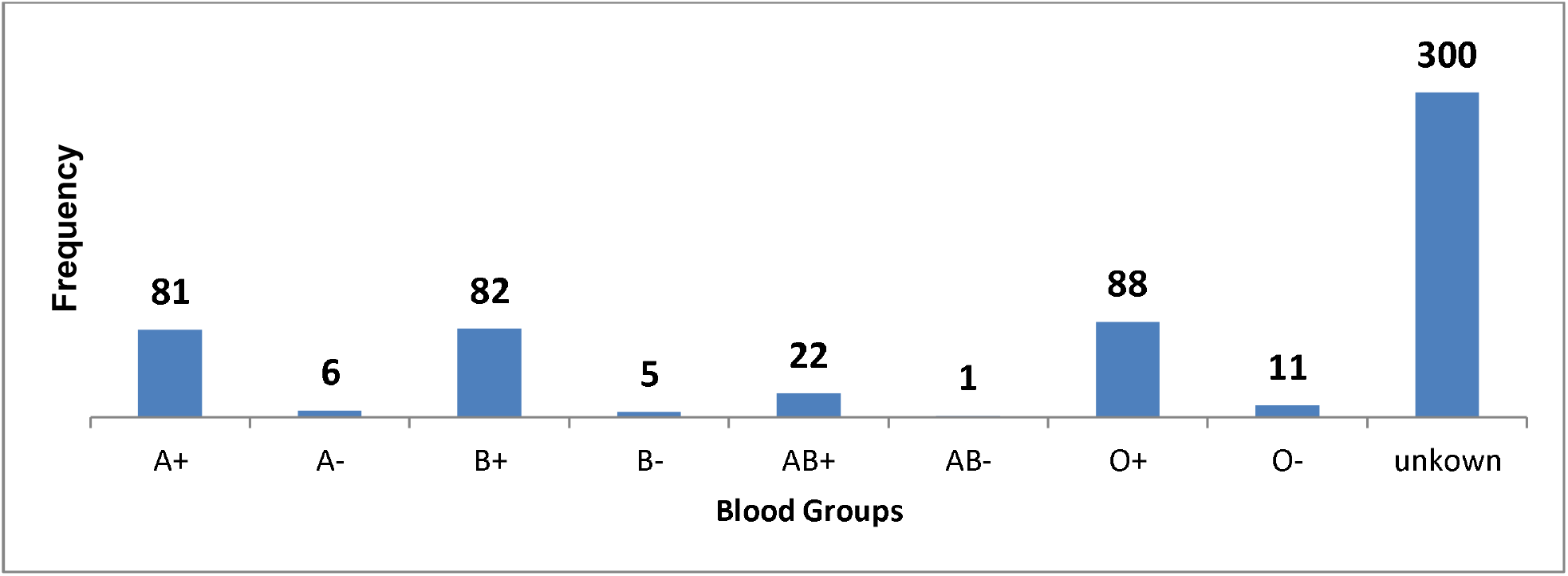
Blood group wise distribution of the study Participants.

### Co morbidities

03% (n=18) participants had Co morbidities while 97% (n=578) had no co morbidity 1.5% (n= 09) had Diabetes Mellitus, 0.16% (n=01) had Asthma, 1.00% (n=06) had HTN, similarly 0.16% (n=01) had Ovarian CA and Hepatitis respectively :

**Fig-5:**
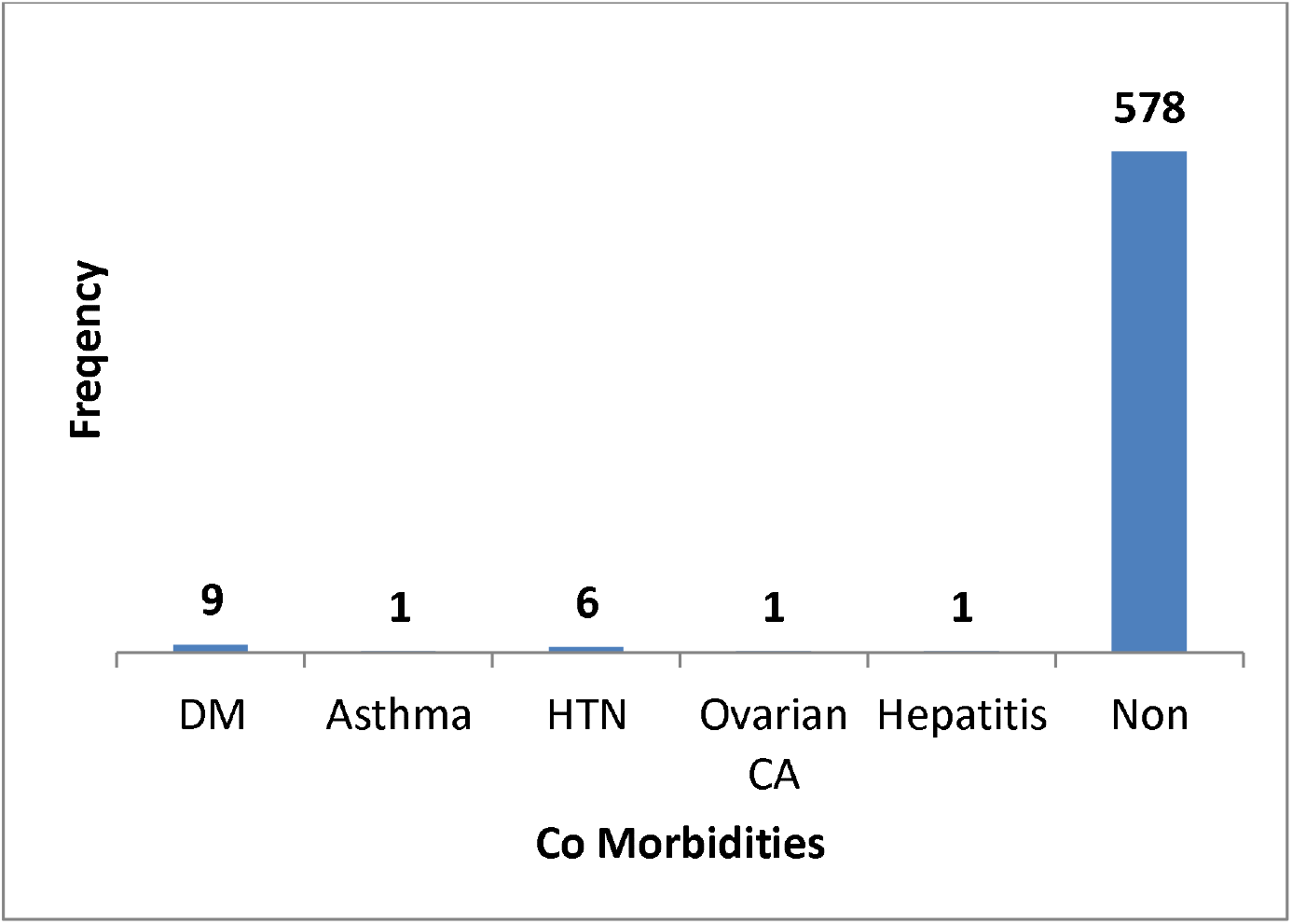
Co Morbidities among the study Participants.

### Occupation

Out of 596 participants only 49% (n=293) were health care providers while 51% (n=303) were non Health Care providers:

**Fig-6:**
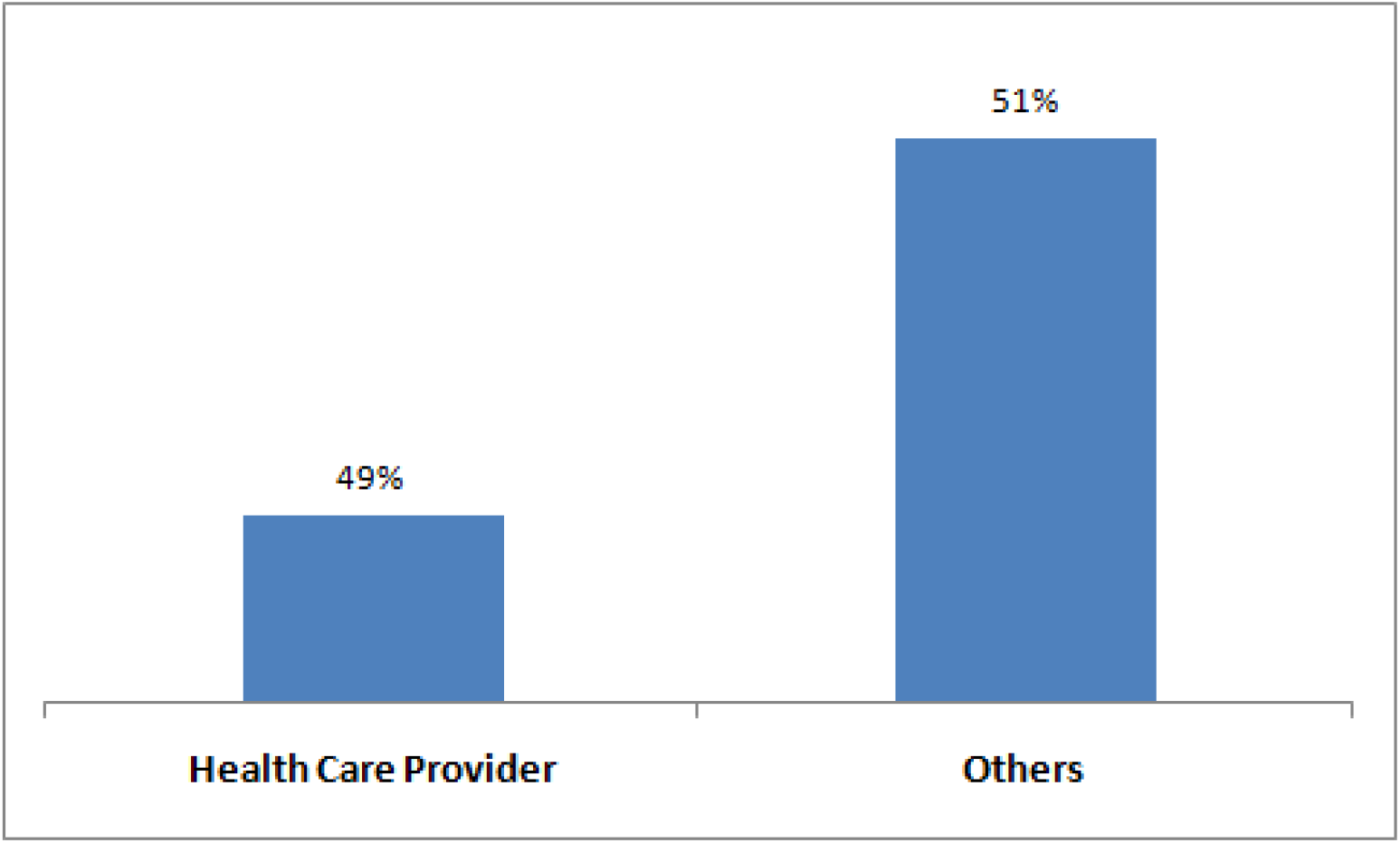
Occupation of the study Participants.

### BCG Vaccination Status

Out of 596 participants 38% (n=229) had never done BCG vaccination while the others 62% (n=367) had done BCG Vaccination:

**Fig-7:**
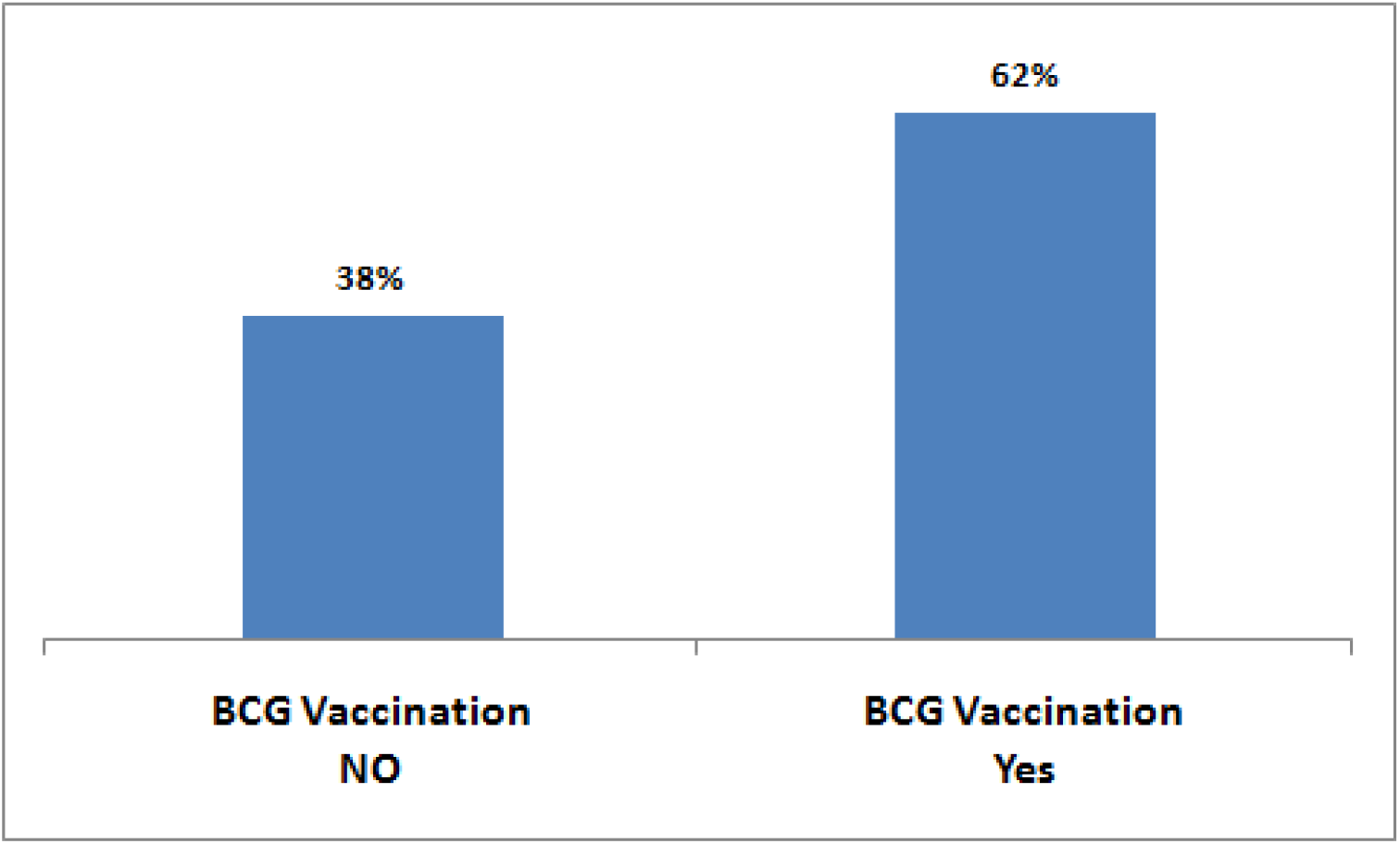
BCG Vaccination Status among the Study Participants.

### Status of Seasonal Flu Vaccination

Out of 596, 07% (n=38) had received seasonal flu vaccination while 93% (n=558) individuals had not received any seasonal flu vaccination.

**Fig-8:**
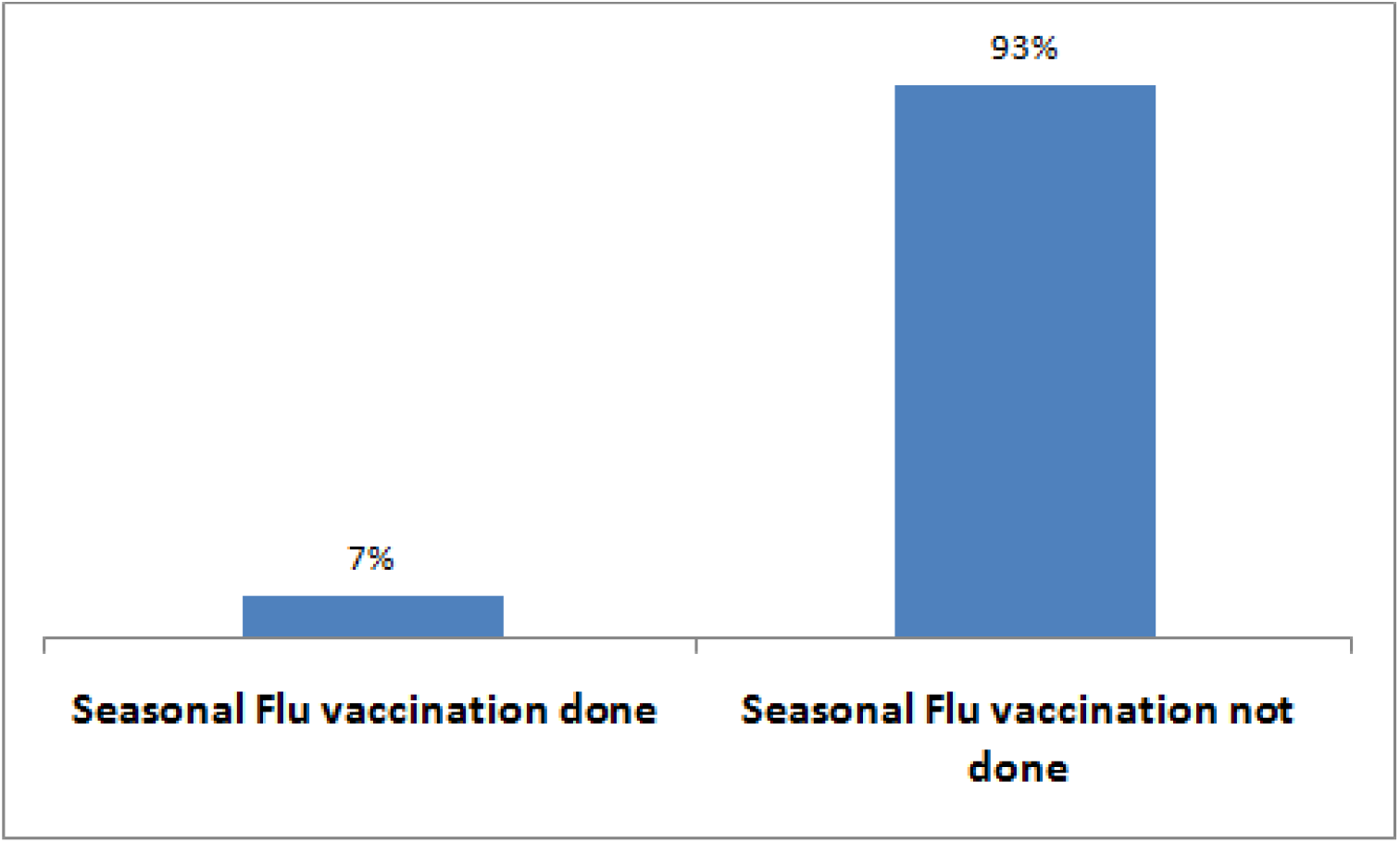
Status of Seasonal Flu among the study Participants.

### History of any close contact or living with confirmed case of COVID-19 in last 15-45 days

Out of 596 participants 59% (n= 351) had contact history during the last 15 to 45 days while 41% (n=245) had no contact history.

**Fig-9:**
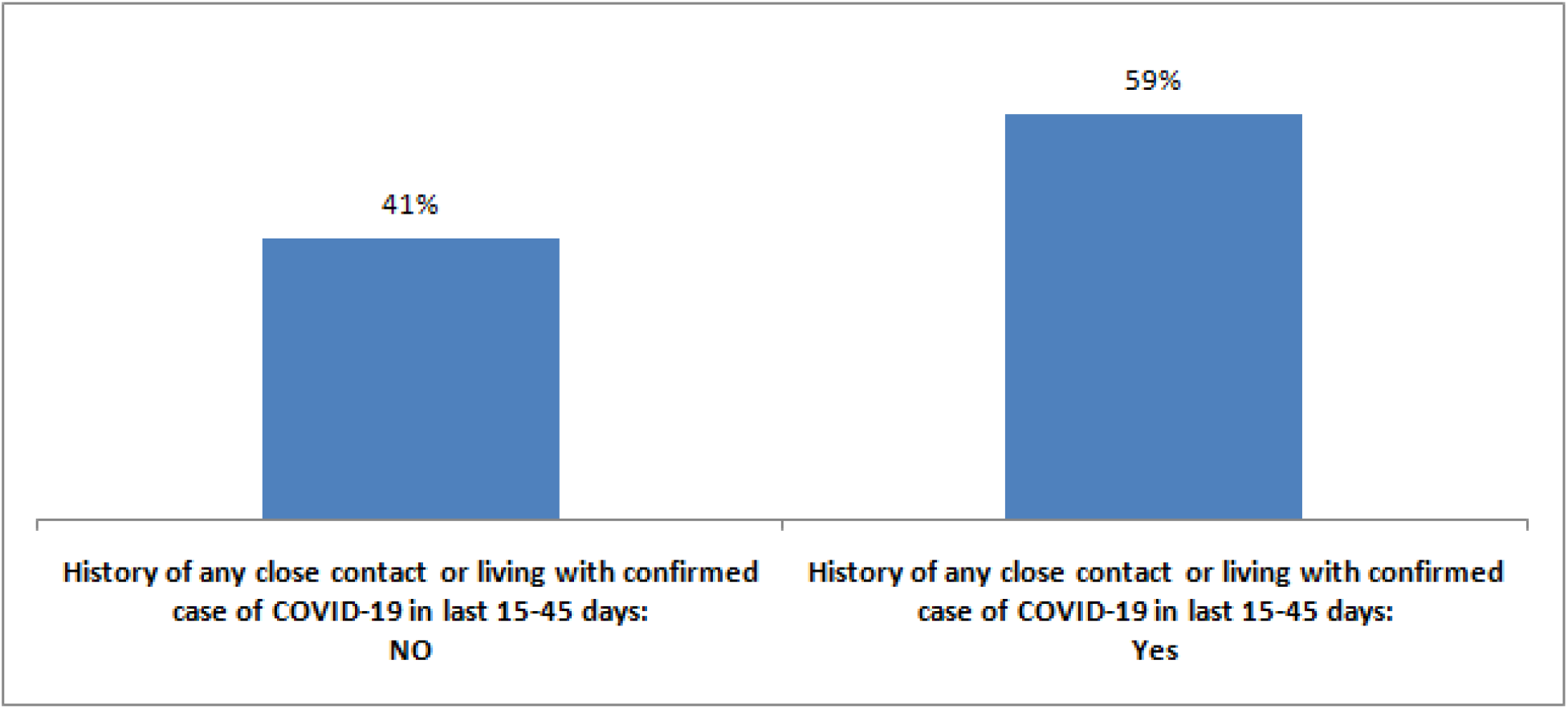
Status of Positive case Contact History.

### Status of Last COVID-19 PCR report

Out of 596 participants 58% (n=346) had negative COVID-19 PCR report while 42% (n=250) had Positive COVID-19 PCR test report:

**Fig 10:**
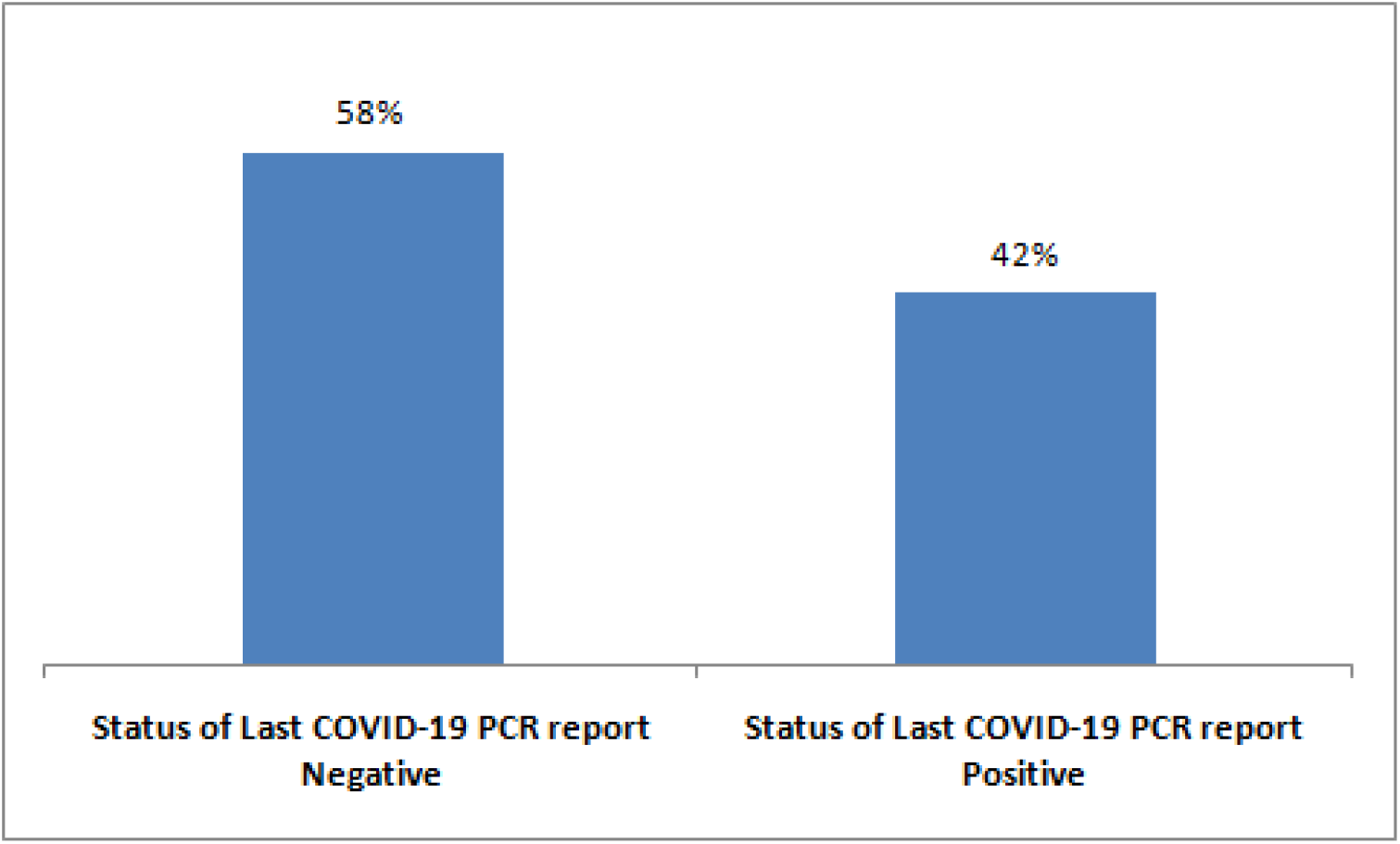
Status of Last COVID-19 PCR Status.

### Rapid Diagnostic Test (RDT) Result status

COVI-19 RDT kits were able to detect antibodies in 49% (n=290) participants; while 51% (n=306) individuals had negative report.

**Fig-11:**
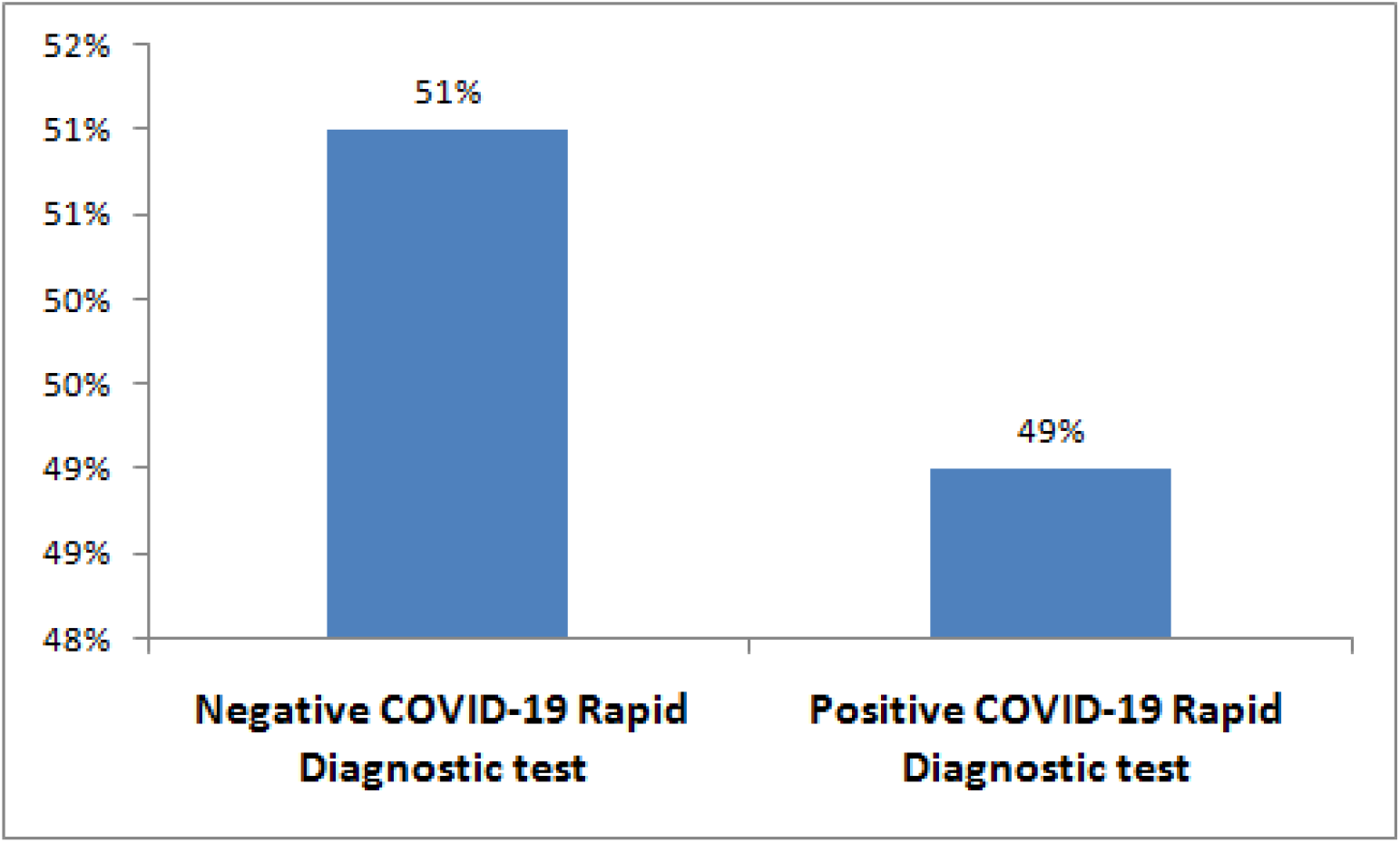
Details of COVID-19 RDT out comes.

### Inferential Statistics

The overall accuracy of the COVID-19 rapid diagnostic test among the study participants was found to be as below:

**Table-1:** 2×2 table for Positive and Negative likely hood ratios calculations.

|  |  | COVID-19 PCR Report |  |  |
| --- | --- | --- | --- | --- |
|  |  | Positive | Negative |  |
| COVID-19<br>RDT TEST<br>Result | POSITIVE | 200 | 90 | 290 |
|  | Negative | 50 | 256 | 306 |
| TOTAL |  | 250 | 346 | 596 |

**Table-2:** Overall Summary sheet. It is clear from the analysis that the Positive Likely hood ratio LR+ is more than 1, while Negative Likely hood ratio is approaching 0. Hence COVID-19 RDT Kits could be used for diagnostic purposes in all Clinical and Screening setup and especially for Test trace and Quarantine (TTQ) activities.

| **When COVID-19**<br>**RDT Tests were Done** | **Sample Size** | **Sensitivity** | **Specificity** | **Positive Likely**<br>**Hood Ratio**<br>**LR+** | **Negative Likely**<br>**Hood Ratio**<br>**LR-** |
| --- | --- | --- | --- | --- | --- |
| They were done<br>after the 14 days of Quarantine<br>& Last PCR Reports. | 596 | 39.20% | 86% | 2.8 | 0.7 |

Likewise Chi-square test of association showed:

**Table-3:** Chi-square test of association. Statistically significant association was found between RDT out comes and Last PCR Test status, Occupation and Contact with COVID-19 positive individuals.

| | RDT Results | | $\chi^2$ (df) | p-value |
| --- | --- | --- | --- | --- |
|  | Positive | Negative |  |  |
| **Last PCR Test Status** |  |  |  |  |
| Positive | **200(34%)** | **50 (8.3%)** | 194.52(1) | 0.00 |
| Negative | **90(15.10%)** | **293(49.1%)** |  |  |
| **Gender** |  |  |  |  |
| Male | **186(31.2%)** | **271(45.5%)** | 3.480(1) | 0.062 |
| Female | **69(11.6%)** | **70(11.7%)** |  |  |
| **BCG Mark of Right Arm** |  |  |  |  |
| Present | **147(24.7%)** | **220(36.9%)** | 2.910(1) | 0.085 |
| Absent | **108(18.1%)** | **121(20.3%)** |  |  |
| **History of Sesonal Influenza vaccination** |  |  |  |  |
| Yes | **18(3%)** | **20(3.4%)** | 0.348(1) | 0.555 |
| No | **237(39.8%)** | **321(53.9%)** |  |  |
| **Occupation-Health Care Provider** |  |  |  |  |
| Yes | **110(18.5%)** | **183(30.7%)** | 6.471(1) | 0.011 |
| No | **145(24.3%)** | **158(26.5%)** |  |  |
| **Close contact or living with confirmed case of COVID-19 in last 15-45 days** |  |  |  |  |
| Yes | **109(18.3%)** | **174(29.2%)** | 4.012(1) | 0.045 |
| No | **146(24.5%)** | **167(28%)** |  |  |

## Discussion

The major findings of this study are almost in line with the set hypothesis and objectives of this study, this study is clearly showing that the Positive Likely hood ratio of the COVID-19 RDT Kits (LR+) is well above 1; similarly the Negative Likely hood ratio is approaching 0.On the other hand the Sensitivity and Specificity 80% and 74% respectively. Similarly study found statistically significant association was between RDT out comes and Last PCR Test status, Occupation and Contact with COVID-19 positive individuals. While other variables like Gender, BCG Vaccination and history of seasonal flu vaccinations were found to have no significant associations with COVID-19 RDT Kit out comes.

The results from this study seems to be consistent with previous studies like in his study Li et al was able to report COVID-19 RDT kits sensitivity as 88.7%, specificity as 90.6%,LR+ = ratio as 9.46 while LR- = ratio as 0.13^(8)^. In another study Ying et al has reported sensitivity to be 85.6%,Specificity as 91.0%,LR+ = 9.52 and LR- = 0.16^(9)^. Which in comparison to this our study has reported _ sensitivity 80%, specificity of 74% respectively while LR+ = 3.07 and LR- = 0.28. All the findings of our study are almost identical. It can be clearly seen that in all three studies COVID-19 RDT Kits (LR+) is well above 1; similarly the Negative Likely hood ratio is approaching 0.

## Conclusion

Being the first study of its kind in Pakistan the major findings of this study are almost in line with the set hypothesis and objectives of this study and based on study findings it will be of high value to use COVID-19 RDT kits during mass screening especially during Test, Trace and Quarantine activities. These kits are reliable, cheap, easy to use and quick to produce results. With such promising likely hood ratios it will be convenient to use these kits for bed site diagnosis of COVID-19.

## Data Availability

will share data file if asked for it.

## Notes

### Competing Interest Statement

The authors have declared no competing interest.

### Funding Statement

No funding was used

### Author Declarations

A local IRB affiliated with Public health Institute of Quetta Balochistan has approved it, the board was chaired by Director General Health services Balochistan.The IRB provided approval on 12/08/2020 and DGHSC-IRB-1234 qta.

